# The Scope and Limitations of Extant Research into ChatGPT as a Tool for Patient Education: Systematic Review

**DOI:** 10.1101/2025.05.20.25328009

**Authors:** Reid Dale, Maggie Cheng, Katharine Casselman Pines, Maria Elizabeth Currie

## Abstract

**Background:** Chat Generative Pre-Trained Transformer (ChatGPT), a large language model (LLM) developed by OpenAI, has been extensively studied and embraced by medical researchers since its public release in November 2022. In addition to its benefits in generating summaries and predictive diagnostics, it has been proposed as a patient education tool. Existing research cites ChatGPT’s potential to increase information availability and accessibility, improve the efficiency of clinical practice, and its high-quality responses to clinical questions as reasons to consider its adoption.

**Objective:** We assessed literature from PubMed on the quality and consistency of ChatGPT responses across medical specialties, evaluated the comprehensiveness of current research, and identified areas for future research.

**Methods:** We searched PubMed for published articles evaluating the consistency, reliability, and ethics of ChatGPT in patient education. Following the PRISMA guideline, we conducted a systematic literature review of the 567 retrieved records. After title, abstract, and full-text screens, 123 relevant records were included and synthesized in this review.

**Results:** We found a lack of consensus among ChatGPT studies. The model accuracy suffers from infrequent updates and generation of misleading information (hallucinations), and it lacks knowledge of current clinical guidelines. The consistency of the model falls short due to its sensitivity to prompt design and fine-tuning through user interaction, making ChatGPT research results almost impossible to peer review and validate. Relying on ChatGPT for clinical information risks spreading misinformation, disrupting trust in the medical system, and disobeying the principles of patient-centered care. This also shifts the focus of patient education from shared decision-making and information-building to information-giving, deviating from the objectives of Health Communication and Health Literacy outlined in Healthy People 2030.

**Conclusions:** We caution against the acceptance of ChatGPT as a patient education tool and encourage future efforts to incorporate community perspectives in AI research.

## Introduction

Chat Generative Pre-Trained Transformer (ChatGPT), initially developed by OpenAI in 2020, is a large language model (LLM) based on GPT-3.5. It was trained with a general information database and can recognize patterns in languages, extract information, and generate coherent and human-like texts.^1–4^ GPT models have undergone several revisions, from GPT-1 in 2018 to GPT-4 in 2023, with progressively increased parameters and training dataset sizes. Although some studies noted significant improvement in accuracy as the model developed (especially comparing GPT 3.5 to GPT 4), others concluded that limitations in previous versions of GPT models still apply to the newest edition now.^4–7^

Although ChatGPT shows excellent performance in sourcing and summarizing information from its training data and could be useful in improving the quality of writing, it is fundamentally a language-based model that lacks expertise in specific subject areas.^3,8–14^ When applied to medical settings as a tool for individualized information sourcing or patient education, its inconsistency and lack of up-to-date medical knowledge raise technical and ethical concerns. Despite the presence of BioGPT, a model trained with biomedical research texts that presumably should be more accurate and rigorous in biomedical settings, the limitations of ChatGPT still apply.^4,15^ At the current stage, although artificial intelligence (AI) technologies have been incorporated into some medical settings such as designing radiology treatment, the incorporation of ChatGPT in offering clinical information to patients remains controversial.^16^

Much of the medical community and published literature support the utilization of ChatGPT in aiding clinical decision-making and as a valuable information source for patients despite recognizing the many flaws this technology carries, citing the model’s near-instantaneous responses, accessibility, affordability, breadth of knowledge, capacity for a personalized experience, and minimization of bias with ongoing efforts to improve training dataset.^4,12,16–41^ However, opposite voices also exist, bringing to light the inconsistency, inaccuracy, privacy, and legal concerns regarding the incorporation of ChatGPT into clinical settings.^2,7,42–58^

In this study, we reviewed literature from PubMed on the design, expandability, consistency, and accuracy of ChatGPT responses across medical specialties, evaluated current research, and identified areas for future research. Despite recognizing ChatGPT’s merits in increasing efficiency and the accessibility of information, it is imperative to critically evaluate the potential concerns in embracing this change too quickly.

## Methods

### Search Strategy

This comprehensive review was conducted according to the Preferred Reporting Items for Systematic Reviews and Meta-Analyses (PRISMA) guidelines. We searched PubMed for published articles (including preprints) on ChatGPT and its applications in providing medical information to patients. Keywords and search terms included “ChatGPT AND consistency”, “ChatGPT AND ethics”, “ChatGPT AND health literacy”, “ChatGPT AND patient education”, and “ChatGPT AND reliability.” Literature search concluded on January 7, 2024.

### Inclusion and Exclusion Criteria

We included published articles (including preprints) on the use of ChatGPT in clinical information dissemination as well as ethics discussion of the model. Exclusion criteria included 1) Letter to Editors (article type), 2) Comments (article type), 3) Records discussing role of ChatGPT in professional/medical education, 4) ChatGPT in scientific writing and publishing, 5) ChatGPT-assisted writing, 6) ChatGPT in diagnostics and imaging, 7) ChatGPT in translation, 8) non-English records, and 9) other irrelevant records. We did not restrict the search and inclusion criteria only to records from the United States. Articles from other countries were included as long as the full text was available and accessible in English.

## Results

### Summary of Search Results and Record Screening Process

A total of 567 records were retrieved from PubMed. These represent a pooled result from all search terms: “ChatGPT AND consistency” (n = 97), “ChatGPT AND ethics” (n = 245), “ChatGPT AND health literacy” (n = 26), “ChatGPT AND patient education” (n = 64), and “ChatGPT AND reliability” (n = 136). 87 records were removed for duplication. A title screen (n = 481) was conducted next, and 274 articles that fitted the exclusion criteria were excluded. Article abstracts (n = 207) were retrieved and screened; 2 were removed for lack of access to the abstract, 14 were removed due to article type, and 56 were excluded for outside of the scope of this review. Lastly, we did a full-text article screen (n = 135); 7 articles were removed for lack of access, 3 were removed for being non-English records, and 1 were removed for being outside the scope of this review. At the end, 123 articles were included in this review. *Figure 1* shows this process in a PRISMA diagram.

**Figure 1.**
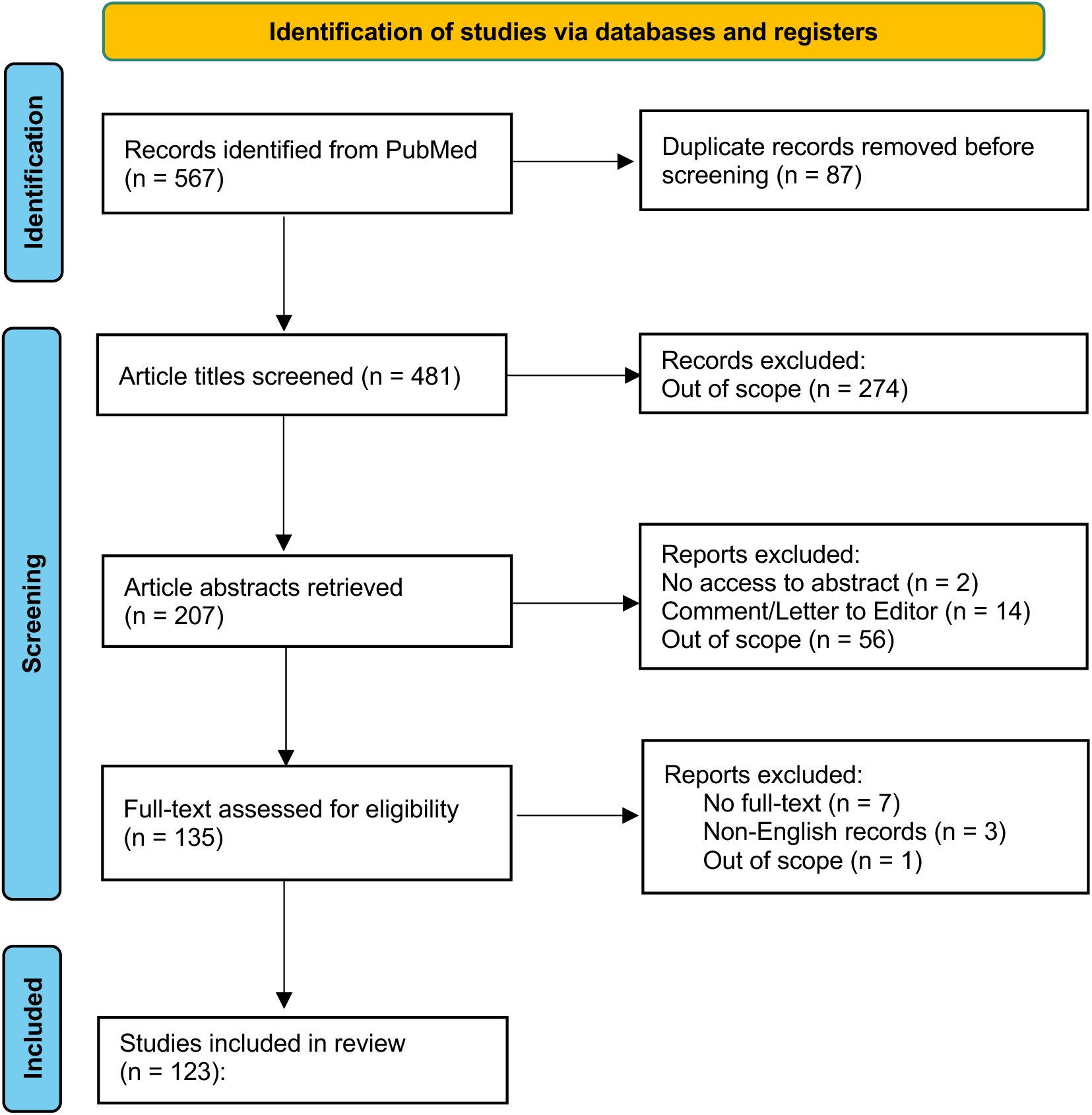
PRISMA 2020 flow diagram for new systematic reviews which included searches of databases and registers only.

### ChatGPT Design

The development of ChatGPT and other ChatGPT-like AI chatbots have the potential to revolutionize some facets of healthcare delivery. Although these models were not developed specifically for a healthcare context, such models could lead to more effective delegation of clinical responsibilities, increase accessibility to health information, and improve clinical decision-making.^3,59^ Beyond sourcing information, ChatGPT also summarizes and presents information concisely, saving readers time from visiting multiple different sites to extrapolate information themselves.^60,61^ It also can translate, making information available and accessible to patients of all language backgrounds, although the accuracy and reliability of ChatGPT in non-English contexts is yet to be fully evaluated.^4,16^ The readability of ChatGPT-generated content has also shown to be above the level that is appropriate to use in educational materials for the general public.^22,50,61–69^

ChatGPT is a pre-trained model with the ability to fine-tune through user interactions. With delays in updating the training dataset and the resources needed for such training, ChatGPT’s response lags behind current clinical guidelines and provides outdated information. GPT 3.5 was updated in January 2022, and GPT 4-Turbo was trained in April 2023.^70^ Updates in clinical guidelines after those dates would be neglected in GPT responses, potentially leading to inaccurate information.^71^ ChatGPT is also subject to temperature control and top_p sampling, which determines the flexibility of the model in selecting the immediate next word as well as the set of phrases to use when generating responses. Lower temperature and top parameters give the model less flexibility in creatively generating responses and higher parameters give the model a higher degree of randomness and unpredictability. As both parameters are non-zero in GPT models, it is almost impossible to replicate an exact same response due to the irreducibly probabilistic nature of an LLM’s response.^72,73^

## Discussion

### Expandability and Privacy Concerns of ChatGPT

Existing studies identified several concerns regarding the usage of ChatGPT in a health education context.

First, the language model operates like a “black box”, not explaining its reasoning or judgments.^16,74,75^ From the user’s perspective, we could only control the input but would not know what to expect as the output. The process that ChatGPT uses to generate responses is unclear, even to its developers from OpenAI.^76^ This takes away the shared nature of decision-making that is supposed to happen in a clinical setting, deviating from the ideals of patient-centered medicine that the medical communities strive to emphasize. Without information on how the model arrives at its recommendations, providers are ill-prepared should patients have any questions about ChatGPT’s responses. Such an inability to properly explain or trace back the logic of AI may instill distrust between the patient and the provider, painting a picture of AI as more knowledgeable while the provider as inferior. Without information on how machine learning models work to generate a response, providers do not have adequate evidence to trust or question the validity of the conclusion, putting them in a dilemma. Their behaviors and clinical judgments may be affected by their willingness to minimize information discrepancy and maintain trust with patients, incentivizing them to side with information from the AI that they may not have full confidence in. In situations where seeking medical information from AI becomes a habit or norm, and patient is faced with conflicting information from the human physician and the AI, the patient would be unsure of how to proceed. Nov et al. gave early evidence of this trend, showing that participants could only correctly distinguish between AI vs. physician-generated responses 49-85.7% of the times, and they tended to trust ChatGPT responses when it comes to low-risk health questions.^77^ When AI-generated information is used and acted upon in clinical settings, the decisions patients make may not be considered truly informed due to the unknown clinical reasoning process of ChatGPT.

Second, medico-legal considerations are widely mentioned in the published literature.^35,47,57,60,78^ To date, we lack regulations and accountability measures that govern the proper utilization of AI-based chatbots in medical settings.^79–83^ Developers are not required to publicly reveal when their models may fail.^16^ In the case of a medical incident, the liability of parties remains unclear.^60,75^ Therefore, when the liability risks are truly at stake, it is unclear whether developers would be as willing to release the AI chatbots to the general public as they do now.

Third, ChatGPT usage brings privacy concerns.^50,57,74^ Nasr et al. revealed possibilities to extract ChatGPT’s training data, raising concerns about confidentiality.^84^ With its ability to widely source and retain information, efforts should be made to limit the possibility of linking de-identified information to personally identifiable information to ensure confidentiality if ChatGPT were to be incorporated in clinical settings.^11^

### Consistency of ChatGPT Responses

With ChatGPT’s sensitivity to prompt design and its constantly evolving nature, mixed results exist regarding the consistency of ChatGPT responses.

Through assessing ChatGPT’s knowledge on five hepato-pancreatico-biliary–related conditions by inputting designed questions into the model three times, Walker et al. showed an internal consistency of 100% of the algorithm.^13^ When two experts compared ChatGPT’s answers with existing guidelines in the study, the interrater agreement showed complete agreement and that the information and diagnoses provided by ChatGPT are consistent with the national guidelines.^13^

However, Johnson et al. found statistically significant differences in the accuracy of ChatGPT responses to their question prompts when the same set of questions were asked to ChatGPT only a few days apart.^85^ Johnson et al. explained such differences as ChatGPT’s ability to learn and fine-tune through user interactions.^85^ However, this can also be interpreted as a lack of consistency in ChatGPT responses. The inconsistency of ChatGPT was further supported by Salas et al. who found that ChatGPT provided different answers to the same prompt within a short time frame.^86^

The finetuning of ChatGPT through user feedback makes it susceptible to manipulation. ChatGPT cannot necessarily distinguish between right or wrong information and lacks factual knowledge in some areas.^87,88^ When ChatGPT is corrected by the users, the corrections could potentially enter the training data of ChatGPT and affect model performance in the future.^87^ If the user feedback were to be incorrect, the model could include inaccurate information or biased views in its training set, allowing the spread of misinformation iteratively.

A recent breakdown of the ChatGPT system raised concerns about the consistency of the model. Deviating from its usual behavior, users reported that the chatbot gave incorrect, irrelevant, and inappropriate answers to their questions and sometimes responded with the same phrase repeatedly without providing a comprehensible sentence. Although this system breakdown was quickly fixed by the developers, OpenAI could not explain why the model failed and admitted that the model behavior can be unpredictable.^76^ Such unpredictability should not be tolerated if the model were to be used for health education or information collection, where each piece of information could inform one’s health decision-making and affect trust in the medical system. With increasingly positive comments from the medical community on ChatGPT’s use in clinics, patients may see those AI chatbots as increasingly reliable, therefore developing dependence on the use of such algorithms and foregoing the knowledge they may gain from interacting with human practitioners.^30,31^

Moreover, ChatGPT is an LLM that detects patterns in language, and it is susceptible to prompt design variations. Jang and Lukasiewicz showed that both ChatGPT (based on GPT-3) and GPT-4 can be self-contradictory, generating different answers when the input questions convey the same meaning.^89^ The same team also demonstrated that GPT-3 often violates logical consistency, and the frequency of such mistakes is not negligible. These limitations were observed in previous versions of GPT models like GPT-2 but remained uncorrected despite using a larger training dataset for subsequent GPT models.

### Accuracy and Comprehensiveness of ChatGPT Responses

When it comes to the accuracy and comprehensiveness of answers, ChatGPT’s responses vary by field. Many studies commented highly on the accuracy of ChatGPT in providing medical information.^85,90–92^ However, events of AI hallucination sometimes appear, providing information or sources that do not exist.^8,13,35,46,87,93,94^ The accuracy of ChatGPT reported by current literature range form 40-50% in some fields (e.g. hepatocellular carcinoma, retinal diseases, liver cancer) to greater than 90% in others (e.g. type 2 diabetes, thoracic surgery, robotic-assisted radical prostatectomy, otolaryngology).^11,25,26,41,95–98^ Assessing ChatGPT’s knowledge across 17 specialties, Johnson et al. found that 8.3% of all answers generated were completely incorrect, and many more were acceptable but not comprehensive. Even for a new GPT model that was trained on biomedical texts, its accuracy was only 51%.^85^ The consistency and accuracy of ChatGPT responses are expected to decrease with increased length of conversation, as ChatGPT sometimes struggles to recognize or maintain context in prolonged conversations.^4,99^

Since ChatGPT is a pre-trained model with limited fine-tuning during use, its accuracy depends on the quality of the training data as well as the frequency of updates. Biases carried in training data will be represented, if not increased, in use.^2,8,47,74,78,100^ Alarmingly, Acerbi and Stubbersfield revealed that ChatGPT-3 showed bias towards content that is gender-stereotypical, negative, and threat-related, warning the public of the limitation of such model.^101^ Koranteng et al. found similar gender stereotypes in ChatGPT responses as well as its tendency to associate negative terms with African American names.^102^ Also, inadequate updates of the model lead to outdated and misleading information.^1,71,103^ Studying ChatGPT’s response to misconceptions about vaccinations, Deiana et al. found various inaccuracies in responses from both ChatGPT GPT 3.5 and GPT 4, and the chatbot seemed to selectively disregard the benefits of vaccines.^1^ Similarly, many other studies across specialties also pointed out that the information ChatGPT provides is not up to date and can be fabricated.^71,91,104,105^

In addition to generating inaccurate answers to the user’s questions, ChatGPT also frequently fails to provide accurate or accessible references, if at all. Eleven of the included articles pointed out that ChatGPT either did not provide references despite prompting or generated fabricated references that did not exist.^93,105–114^ This inability to cite its sources exist regardless of the information accuracy provided by ChatGPT. Alessandri-Bonetti et al. showed that among all requests, ChatGPT failed to provide references 33% of the time.^106^ When references were provided, 36% were shown to be irrelevant or nonexistent.^106^ McGowan et al. revealed that only 6% of the references provided by ChatGPT in their study was accurate.^105^ ChatGPT also has limited ability to assess the quality of such citations, potentially biasing towards sources that are present in higher quantity in its training data but not necessarily of high quality or accuracy.

### Potential Impacts on Patient-Physician Relationship

Although only a few of the current studies clearly advice the incorporation of ChatGPT in clinical setting for information retrieval purposes, much of the literature supports the idea of using ChatGPT as a supplementary tool to aid health professionals in promoting patient education and health literacy.^11,60,115–122^ Whereas the efficiency and relative accuracy of ChatGPT maybe appealing, we should not overlook the challenges ChatGPT faces as well as the patient-provider relationship that is vital in building health literacy.^123,124^ Health literacy must not only advance people’s knowledge and ability to obtain and understand health information, but also empower them to actively participate in the health decision-making process. This includes discerning the quality of information, communicating their concerns, asking questions, and self-manage their conditions.^16,123–126^

Putting the accuracy and consistency concerns about ChatGPT aside, merely providing information to patients is not an effective way to improve health literacy and can be dangerously biased. Just as the black-box nature of GPT models goes against the key principles of patient-centered medicine and informed consent, receiving health advice based on the limited patient-provided information to ChatGPT carries the same risks. Modern medicine emphasizes both evidence-based practice and patient-centered care. Recommending the utilization of ChatGPT for obtaining medical information may dangerously reduce people into numbers and data points and potentially ignore their beliefs and values.^16^ On the contrary, healthcare professionals play a key role in incorporating such considerations through the establishment of a trusting patient-provider relationship.^16^ Such a relationship should not be a hierarchical one of information-giving and receiving or one where providers work to correct recommendation inaccuracies from AI-based chatbots. Instead, it should be one of information- and relationship-building where the patient takes a proactive part to work with the healthcare provider and arrive at a final decision. ChatGPT also lacks incentives to improve its performance standards. Human practitioners have moral obligations and liability of malpractice and negligence to provide high standards of care, while this ability is not yet developed in ChatGPT.

Since ChatGPT is sensitive to prompt design and has the potential to provide irrelevant information, utilizing ChatGPT properly and reliably requires knowledge and skills to ask the right questions and assess the quality of the information received that not everyone possesses.^16,67,126–128^ Although ChatGPT-based information is accessible, it can worsen the existing disparities around health literacy due to this initial knowledge threshold required to use the model. If not done correctly, ChatGPT may miss important information, give generic answers that are not specific to the patient’s situation, or provide misleading or overly worrisome messages.^1,129–131^ As mentioned above, ChatGPT is subject to biases in its training data and memory, both are aspects the patient may not be aware of when obtaining or acting on the medical information provided by ChatGPT.

Although ChatGPT has an excellent ability to generate human-like and coherent texts, it cannot think like a medical professional or ask follow-up questions. On the contrary, medical professionals are crucial not only hearing the patients’ experiences, but also offering clarification, actively engaging patients to ensure proper understanding, and providing emotional support.

Therefore, if ChatGPT or any AI-based sources were to be implemented in patient education in the future, close monitoring and guidance by medical professionals would be necessary to help patients develop the ability to identify reliable and authoritative sources and critically assess the quality of information online before they can be left on their own. These skills are especially important to develop with the increased democratization of knowledge and autonomy patients have in their own health education.^1,85^

### Gaps in ChatGPT Research

Primary studies that evaluate the consistency and quality of ChatGPT’s responses vary in rigorousness. Some studies rate ChatGPT’s responses on a Likert scale while others report the accuracy of ChatGPT responses in percentages. The lack of standardized metrics makes it hard to compare the accuracy of the model across specialties.

Mixed results exist in many areas of ChatGPT research. Some showed an internal consistency of 100% and commented highly on its accuracy while others found that even the most updated GPT model (GPT-4) frequently self-contradict, and ChatGPT only has an accuracy of around or less than 50%.^11,13^

The sensitivity of the model to prompt design, its memory bias, the temperature control, and its constantly evolving nature make it almost impossible to peer review, reproduce, and validate study findings around ChatGPT. Contradicting results across studies can be justified as ChatGPT’s ability to learn and evolve, while it may also be interpreted as a sign of inconsistency in the ChatGPT model.

Many other studies compare ChatGPT to Google searches or other AI-based chatbots (Google Bard and BingAI), stating that ChatGPT’s responses are comparable to or more valuable than the ones provided by Google and other AI chatbots.^28,62,63,66,106,132–135^ However, this comparison does not that ChatGPT should be used and trusted for medical information.

Furthermore, current ChatGPT research lacks a community perspective, using expert-designed or guideline-oriented questions to generate ChatGPT responses and seeking experts’ views on incorporating this in clinical practice. Although healthcare professionals play a crucial role in clinical interactions, they are only part of the story. Patients’ perspectives should also be gathered and incorporated through community outreach efforts. With ChatGPT’s sensitivity to instructions and prompt design, the differences in questions asked between professionals and a layperson without much background in a specific topic area can make a significant difference in the quality of response they get.

### Future Directions

The crucial community perspective is lacking in current ChatGPT research. Only two studies among all that are included in this review sought community’s expertise on this issue.^77,136^ A crucial next step is to continue this effort, survey patients, and evaluate their experience, trust, and comfortability in utilizing AI-based models for medical information.^137^ Although comparison between different versions of GPT models did not reach a consensus, there is early evidence suggesting a superior performance in the paid GPT 4 compared to the publicly accessible ChatGPT (GPT 3.5). This raises concerns from a health equity standpoint, and future research is needed to address this issue.

Inadequacies of ChatGPT are not unique to the model and can extend to other AI-based chatbots as well. With Nvidia and Hippocratic AI’s recent collaboration on an empathetic AI agent that outperformed human nurses in offering clinical information and decisions, the speed and scope of such technological developments have been pushed to a new height, making accountability and ethical evaluations of such practices especially pressing and necessary.^138^ Although there are some regulations and quality assessment tools in development, such progress is far from catching up with how fast AI-based chatbots has been spread and utilized in daily, academic, and medical settings.^36^ Future research should focus on increasing patients’ awareness of the limitations of AI-based chatbots while establishing regulations to ensure ethical and equitable use of such models.

## Conclusions

We evaluated the potential of incorporating ChatGPT in clinical practice and revealed the inconsistency, inaccuracy, and lack of accountability in the ChatGPT model. With significant delays in model training, events of hallucination, and a lack of transparency in its reasoning, ChatGPT is not yet a reliable source of health information. The purpose of patient education and increasing health literacy involves trust- and relationship-building between the patient and the provider, and it exists beyond what ChatGPT can provide. Therefore, we caution against the premature acceptance of ChatGPT as a patient education tool until its logical, legal, and accuracy concerns are addressed. We encourage future efforts to increase transparency of model development and reasoning and incorporate community perspectives in AI research.

## Data Availability

All data produced in the present study are available upon reasonable request to the authors

## Ethics approval and consent to participate

This study is a review of published literature and is exempt from human subjects research review by the Stanford University Institutional Review Board.

## Availability of data and materials

Not applicable (this manuscript does not report data generation or analysis).

## Conflicts of Interest

The authors declare that they have no competing interests.

## Acknowledgement

Dr. Reid Dale is supported by Stanford University Department of Cardiothoracic Surgery [grant number 1252717-910-EAGHD].

## Authors’ contributions

Framework and design of the review: R. D. and M, E. C.; literature searches and review: M. C.; drafting the manuscript: M. C. and R. D.; revision of the manuscript: M. C., K. C. P., R. D., and M. E. C.; final approval of the manuscript for publication: M. C., K. C. P., R. D., and M. E. C.

## List of Abbreviations

AI: Artificial Intelligence
ChatGPT: Chat Generative Pre-Trained Transformer
GPT: Generative Pre-Trained Transformers
LLM: Large Language Model
PRISMA: Preferred Reporting Items for Systematic Reviews and Meta-Analyses

## Notes

### Competing Interest Statement

The authors have declared no competing interest.

